# Compassionate use of recombinant human IL-7-hyFc as a salvage treatment for restoring lymphopenia in patients with recurrent glioblastoma

**DOI:** 10.1101/2022.01.09.22268651

**Authors:** Stephen Ahn, Jae-Sung Park, Heewon Kim, Minkyu Heo, Young Chul Sung, Sin-Soo Jeun

## Abstract

**Purpose:** Lymphopenia is frequently observed and is associated with poor prognosis in glioblastoma (GBM) patients. Restoring lymphopenia in cancer patients has been suggested as a novel immunotherapeutic strategy. As interleukin-7 (IL-7) is necessary for proliferation of lymphocytes and to amplify the total lymphocyte count (TLC), IL-7 therapy has been tried for various cancers, although the results are inconclusive. Here, we describe the clinical results of recurrent GBM treated with long-acting engineered version of recombinant human IL-7 (rhIL-7-hyFc).

**Methods:** This prospective case series based on compassionate use was approved by the Ministry of Food and Drug Safety in South Korea. Patients with recurrent GBM were enrolled to Seoul St. Mary’s Hospital. Primary outcomes were the safety profile and elevated total lymphocyte count (TLC). Secondary outcomes were overall survival (OS) and progression-free survival (PFS). The duration of median follow-up was 372.6 days (range 98–864 days).

**Results:** Among 18 patients enrolled, 10 received rhIL-7-hyFc with temozolomide, 5 received rhIL-7-hyFc with bevacizumab, 1 received rhIL-7-hyFc with PCV chemotherapy, and 2 received rhIL-7-hyFc alone. The mean TLC of enrolled patients after the first treatment with rhIL-7-hyFc was significantly increased from 1,131 cells/mm^3^ (range 330-2,989) at baseline to 4,356 cells/mm^3^ (range 661-22,661). Similar increase was observed in 16 of 18 patients (88.8%), only after the first treatment of rhIL-7-hyFc. TLCs of these patients were maintained higher while rhIL-7-hyFc was repeatedly administered. Most common adverse events were injection sites reactions (64.7%) including urticaria and itching sensation, however, there were no serious adverse events more than grade III. Median OS and PFS were 378 days (range 107-864 days) and 231 days (55-726 days), respectively.

**Conclusion:** Our study first reports that IL-7 immunotherapy can restore lymphopenia and maintain TLC with various salvageable chemotherapies in recurrent GBM patients without serious adverse toxicities. This outcome warrants further larger and randomized clinical trials to validate the clinical benefits of rhIL-7-hyFc for GBM patients.

## Introduction

Glioblastoma is the most common and devastating brain malignancy in adults ^1^. Median overall survival (OS) is less than 2 years despite aggressive multimodal standard of care including maximal safe resection and concomitant chemoradiation (CCRT) followed by adjuvant chemotherapy with temozolomide ^2^. Tumor recurrence and progression occur in almost all GBM patients, in whom available treatment options will be re-surgical resection; re-irradiation; and chemotherapies such as temozolomide, bevacizumab, and PCV [procarbazine, lomustine (CCNU), and vincristine]. However, the clinical benefit is unsatisfactory ^3 4^.

Lymphopenia, defined as decreased counts of circulating lymphocytes resulting from mainly radiotherapy, is frequently observed in GBM patients and is recently established as a novel biomarker associated with poor clinical outcomes ^5 6 7 8 9^. Furthermore, restoring lymphopenia is a novel therapeutic approach in various cancers including GBM ^10 11^. Interleukin-7 (IL-7), which is one of the common gamma-chain cytokines with IL-2 and IL-15 and has critical roles in maintaining lymphocyte homeostasis, is suggested to restore lymphopenia and improve clinical outcomes of cancer patients ^12 13^. In this background, IL-7 cytokine therapy was tested to treat recurrent or refractory cancer patients in clinical studies, but showed unclear efficacy of T cell regeneration and clinical benefits ^14,15, 16, 17^.

In our compassionate use program, our study investigated clinical benefits from the treatment of hybrid-Fc fused recombinant human IL-7 (rhIL-7-hyFc) in recurrent GBM patients. We evaluated whether rhIL-7-hyFc therapy could restore lymphopenia and increase total lymphocyte count (TLC) with salvageable chemotherapies for GBM patients. In addition, we evaluated the safety profiles and survival outcomes of enrolled patients.

## Patients and Methods

### Ethical consideration

Patients with recurrent GBM received rhIL-7-hyFc on compassionate use basis, approved and supervised by the Ministry of Food and Drug Safety in South Korea. The study was conducted according to the Declaration of Helsinki and was approved by the institutional review board in Seoul St. Mary’s Hospital. It was registered in the National Institutes of Health Clinical Trial Registry (NCT04289155). All patients signed on informed consent forms prior to compassionate use of rhIL-7-hyFc.

### Study population

Patients who were pathologically confirmed with recurrent GBM between March 2018 and December 2019 at Seoul St. Mary’s Hospital were screened. Treatment with rhIL7-hyFc was offered for patients older than 18 years with pathologically confirmed recurrent GBM. Patients who had acute infection, autoimmune or hematologic disease at the time of recurrence were excluded.

### Clinical variables

Baseline characteristics of sex, date of birth, dates of surgeries, pathological findings, prior treatments, and radiological findings were collected and summarized. Pathological evaluation was performed by a neuropathologist following the 2016 WHO classification of the central nervous system. Isocitrate dehydrogenase (IDH)1 or IDH2 mutation was assessed by Sanger sequencing. Co-deletion of 1p19q was identified by fluorescent in situ hybridization. [6]-Methylguanine-DNA methyltransferase (MGMT) gene methylation and TERT promoter mutation status were evaluated by polymerase chain reaction (PCR). Loss of ATRX was assessed using immunohistochemistry. Radiographic responses on magnetic resonance imaging (MRI) were determined by two neuro-radiologists according to Immunotherapy Response Assessment in Neuro-Oncology (iRANO) criteria. The duration of recurrence was defined as days from initial surgery to date of MRI showing recurrence.

### Treatment protocols of rhIL-7-hyFc therapy

rhIL-7-hyFc was prepared and provided by Genexine, Inc. and detailed information for rhIL-7-hyFc was previously reported ^18^. In brief, rhIL-7-hyFc is a recombinant human IL-7 fused to the hybridizing IgD/IgG4 immunoglobulin domain to increase half-life of IL-7 *in vivo*. rhIL-7-hyFc was administered intramuscularly into the gluteus muscle and/or deltoid muscle every 4 to 8 weeks. Patients were treated at various dose ranging from 60 μg/kg to 1,440 μg/kg while being monitored for toxicity.

Patients received salvageable systemic therapy with rhIL-7-hyFc treatment, if satisfied with criteria. Systemic therapy regimen was determined by a treating clinician. Temozolomide was used for most recurrent cases occurring > 6 months upon completion of standard of care. Bevacizumab and chemotherapy with procarbazine, lomustine and vincristine (PCV) were allowed depending on clinician’s assessment. Dosing schedule of rhIL-7-hyFc was determined in consideration of both the lympho-depletion effect of cytotoxic chemotherapies and the recovery time of lymphocyte counts induced by rhIL-7-hyFc. In combination treatment with temozolomide, patients were given rhIL-7-hyFc one week after initiation of temozolomide. In combination treatment with bevacizumab, the patients were given rhIL-7-hyFc within 3 days of bevacizumab. In combination therapy with PCV chemotherapy, rhIL-7-hyFc was administered one week after completion of PCV.

### Primary and secondary endpoints

Primary endpoints were safety events and restoration of total lymphocyte count. Treatment-associated toxicity was evaluated at every visit using Common Terminology Criteria for Adverse Events (CTCAE) version 5.0. Complete blood count including white blood cells and composition of neutrophils and lymphocytes were calculated at the time of blood sampling on the day of chemotherapy, day of administration of rhIL-7-hyFc, and 3 weeks after initial administration of rhIL-7-hyFc. Secondary endpoints were overall survival (OS) and progression-free survival (PFS) after recurrence. OS was also defined as days from the date of MRI showing recurrence before use of rhIL-7-hyFc to date of death. Patients alive on February 28, 2021, were censored. The mean duration of follow-up was 372.6 days (range 98–864). To compare the OS of enrolled patients with historical patients; 123 patients with recurrent GBM diagnosed and treated in this hospital between 2014 and 2017 were enrolled into the historical cohort. The detailed characteristics of the historical cohort are described in Supplementary Table 1. These patients were followed by routine magnetic resonance imaging (MRI) every 8 or 12 weeks if they do not exhibit neurologic symptoms.

### Statistical analysis

Continuous clinical variables such as TLC are expressed as mean value ± standard deviation. Swimmer plot and all figures were constructed using GraphPad Prism software (Version 8.4.3). Kaplan–Meier survival was used to estimate median OS and PFS. Log-rank test was used to compare OS between the treatment group and historical control. Statistical analysis was estimated using R Statistical Software (Version 4.0.5).

## Results

### Characteristics and clinical follow-up of patients treated with rhIL-7-hyFc

A total of 18 patients with pathologically confirmed recurrent GBM were included in the study. Clinical characteristics of sex; age; pathological findings; the presence of leptomeningeal spread at enrollment; and history of treatment including surgery, chemotherapy, and radiotherapy before enrollment are described in Table 1. Of 18 patients, 10 started rhIL-7-hyFc treatment with combination of temozolomide chemotherapy (2 patients switched to bevacizumab after progression), 5 received it with combination of bevacizumab with or without irinotecan, 1 received it with PCV chemotherapy, and 2 received rhIL-7 treatment alone. Of these, 72% of patients (13 of 18) received rhIL-7-hyFc at least twice, while 28% of patients (5 of 18) received it only one time. The initial dose was 60 μg/kg and increased to 1,440 μg/kg. Detailed information on rhIL-7-hyFc injection with systemic therapy is described in Table 2, and clinical follow-up of the patients is illustrated in Figure 1.

**Table 1.**
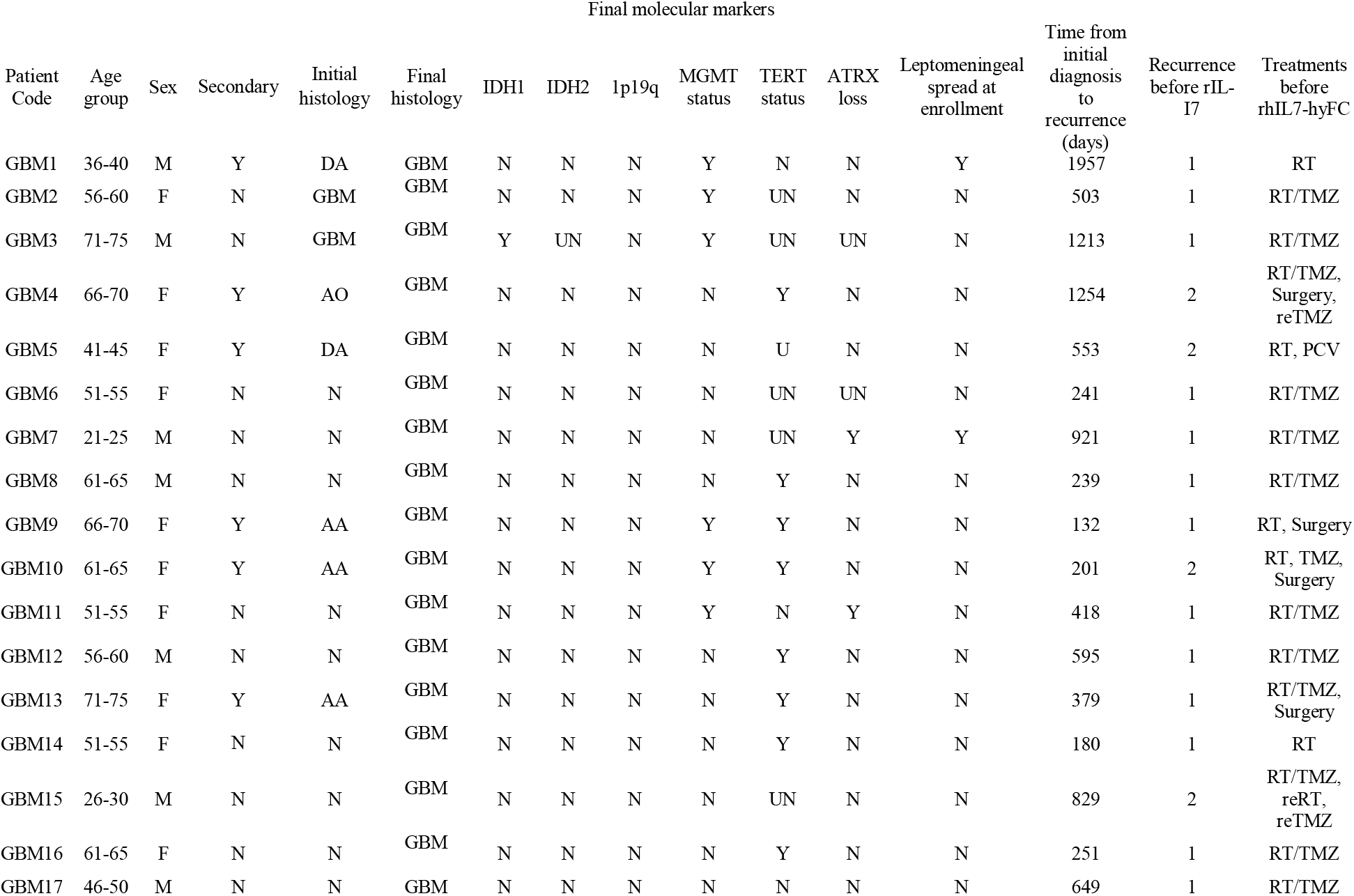
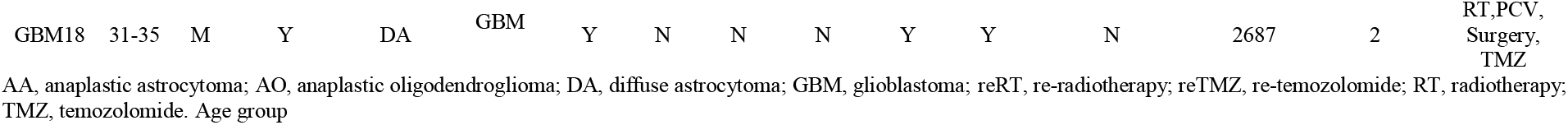
Clinical characteristics of patients with recurrent GBM.

| Patient Code | Age group | Sex | Secondary | Initial histology | Final histology | IDH1 | IDH2 | 1p19q | MGMT status | TERT status | ATRX loss | Leptomeningeal spread at enrollment | Time from initial diagnosis to recurrence (days) | Recurrence before rIL-17 | Treatments before rhIL7-hyFC |
| --- | --- | --- | --- | --- | --- | --- | --- | --- | --- | --- | --- | --- | --- | --- | --- |
| GBM1 | 36-40 | M | Y | DA | GBM | N | N | N | Y | N | N | Y | 1957 | 1 | RT |
| GBM2 | 56-60 | F | N | GBM | GBM | N | N | N | Y | UN | N | N | 503 | 1 | RT/TMZ |
| GBM3 | 71-75 | M | N | GBM | GBM | Y | UN | N | Y | UN | UN | N | 1213 | 1 | RT/TMZ |
| GBM4 | 66-70 | F | Y | AO | GBM | N | N | N | N | Y | N | N | 1254 | 2 | RT/TMZ, Surgery, reTMZ |
| GBM5 | 41-45 | F | Y | DA | GBM | N | N | N | N | U | N | N | 553 | 2 | RT, PCV |
| GBM6 | 51-55 | F | N | N | GBM | N | N | N | N | UN | UN | N | 241 | 1 | RT/TMZ |
| GBM7 | 21-25 | M | N | N | GBM | N | N | N | N | UN | Y | Y | 921 | 1 | RT/TMZ |
| GBM8 | 61-65 | M | N | N | GBM | N | N | N | N | Y | N | N | 239 | 1 | RT/TMZ |
| GBM9 | 66-70 | F | Y | AA | GBM | N | N | N | Y | Y | N | N | 132 | 1 | RT, Surgery |
| GBM10 | 61-65 | F | Y | AA | GBM | N | N | N | Y | Y | N | N | 201 | 2 | RT, TMZ, Surgery |
| GBM11 | 51-55 | F | N | N | GBM | N | N | N | Y | N | Y | N | 418 | 1 | RT/TMZ |
| GBM12 | 56-60 | M | N | N | GBM | N | N | N | N | Y | N | N | 595 | 1 | RT/TMZ |
| GBM13 | 71-75 | F | Y | AA | GBM | N | N | N | N | Y | N | N | 379 | 1 | RT/TMZ, Surgery |
| GBM14 | 51-55 | F | N | N | GBM | N | N | N | N | Y | N | N | 180 | 1 | RT |
| GBM15 | 26-30 | M | N | N | GBM | N | N | N | N | UN | N | N | 829 | 2 | RT/TMZ, reRT, reTMZ |
| GBM16 | 61-65 | F | N | N | GBM | N | N | N | N | Y | N | N | 251 | 1 | RT/TMZ |
| GBM17 | 46-50 | M | N | N | GBM | N | N | N | N | N | N | N | 649 | 1 | RT/TMZ |
| GBM18 | 31-35 | M | Y | DA | GBM | Y | N | N | N | Y | Y | N | 2687 | 2 | RT,PCV,<br>Surgery,<br>TMZ |

AA, anaplastic astrocytoma; AO, anaplastic oligodendroglioma; DA, diffuse astrocytoma; GBM, glioblastoma; reRT, re-radiotherapy; reTMZ, re-temozolomide; RT, radiotherapy; TMZ, temozolomide. Age group

**Table 2.**
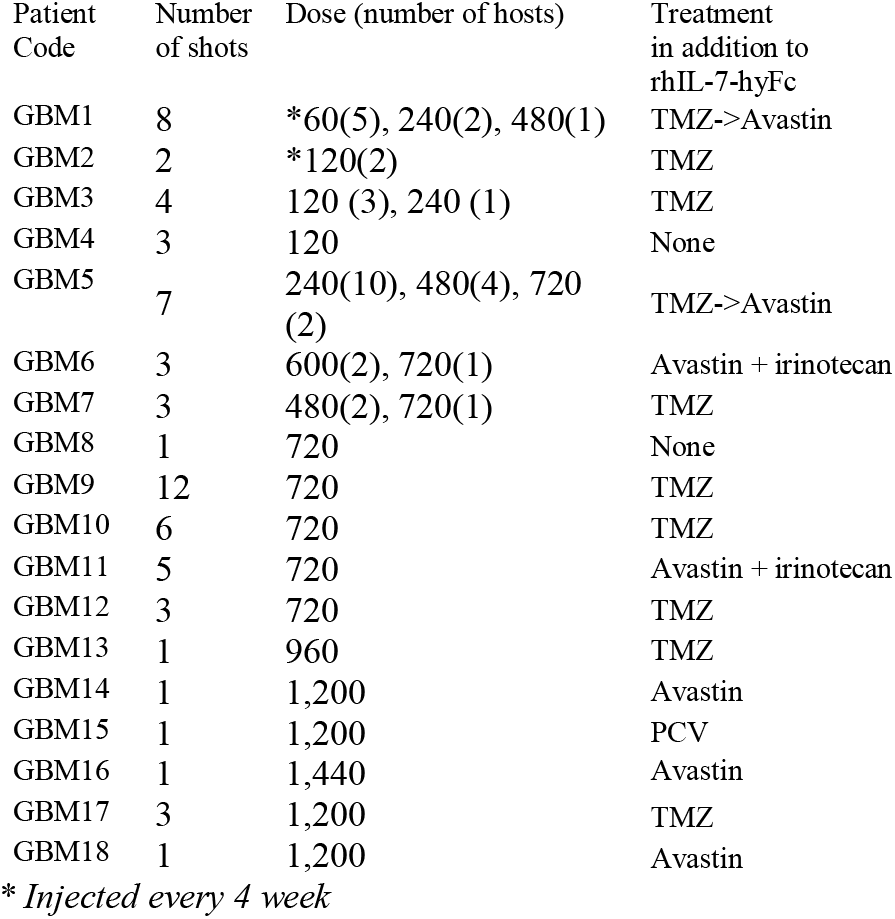
Details of treatment with rhIL-7-hyFc and clinical follow-up of patients with recurrent GBM.

**Figure 1.**
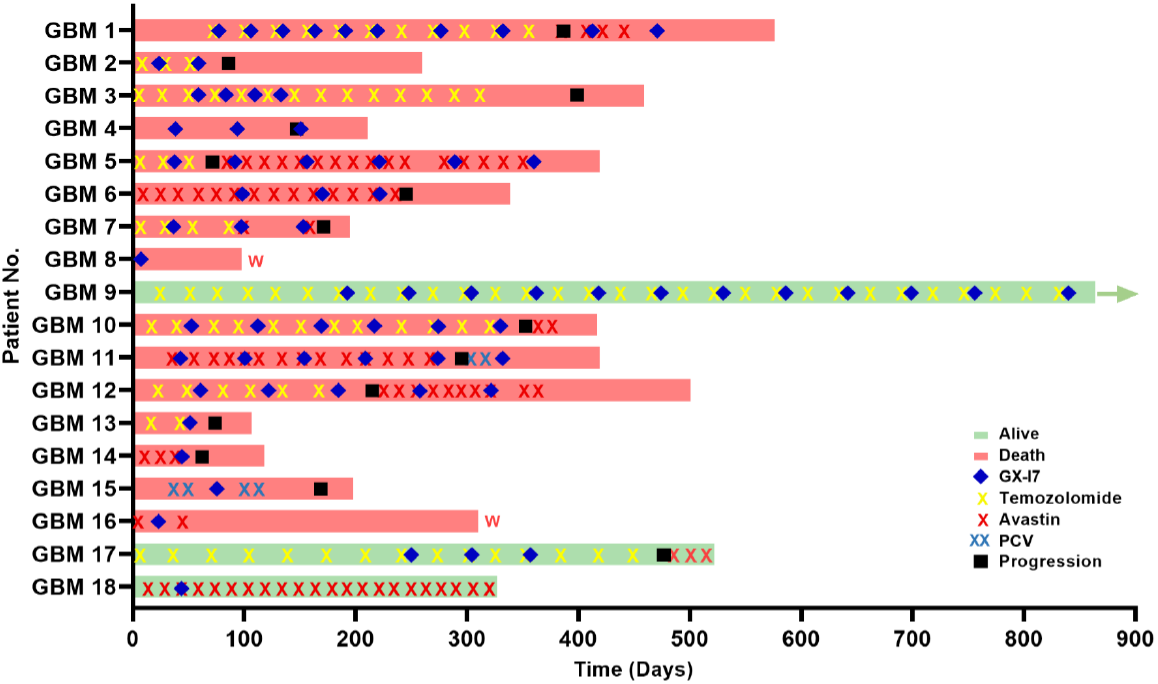
Swimmer plot of clinical course of patients after enrollment. A total of 18 patients were assessed in this program. Each bar represents an individual patient’s treatment history with subsequent treatment and the bar color indicating survival status. These included the following: rhIL-7-hyFc with temozolomide, n = 10; rhIL-7-hyFc with bevacizumab, n = 5; rhIL-7-hyFc with PCV chemotherapy, n = 1; rhIL-7-hyFc alone, n = 2).

### TLC changes following treatment with rhIL-7-hyFc

The mean TLC of these patients before treatment with rhIL-7-hyFc was 1,131 (range 330-2,989) cells/mm^3^. After initial treatment with rhIL-7-hyFc, mean TLC increased to 4,356 (range 661-22,661) cells/mm^3^; a mean 3.40 (range 1.38-9.13)-fold increase. Increased TLC was noted in 16 of 18 patients (94.4%) after the first treatment with rhIL-7-hyFc and it was maintained following repeated treatment; only one patient (GBM 17) did not respond to rhIL-7-hyFc. Changes of TLC in all enrolled patients following injection of rhIL-7-hyFc are described in Supplementary Table 2. The dose of rhIL-7-hyFc was categorized as low (60-240 μg/kg), medium (480-720 μg/kg), and high (960-1,440 μg/kg),; TLC significantly increased in a dose-dependent manner for all patients but one patient (GBM17), as presented in Figure 2.

**Figure 2.**
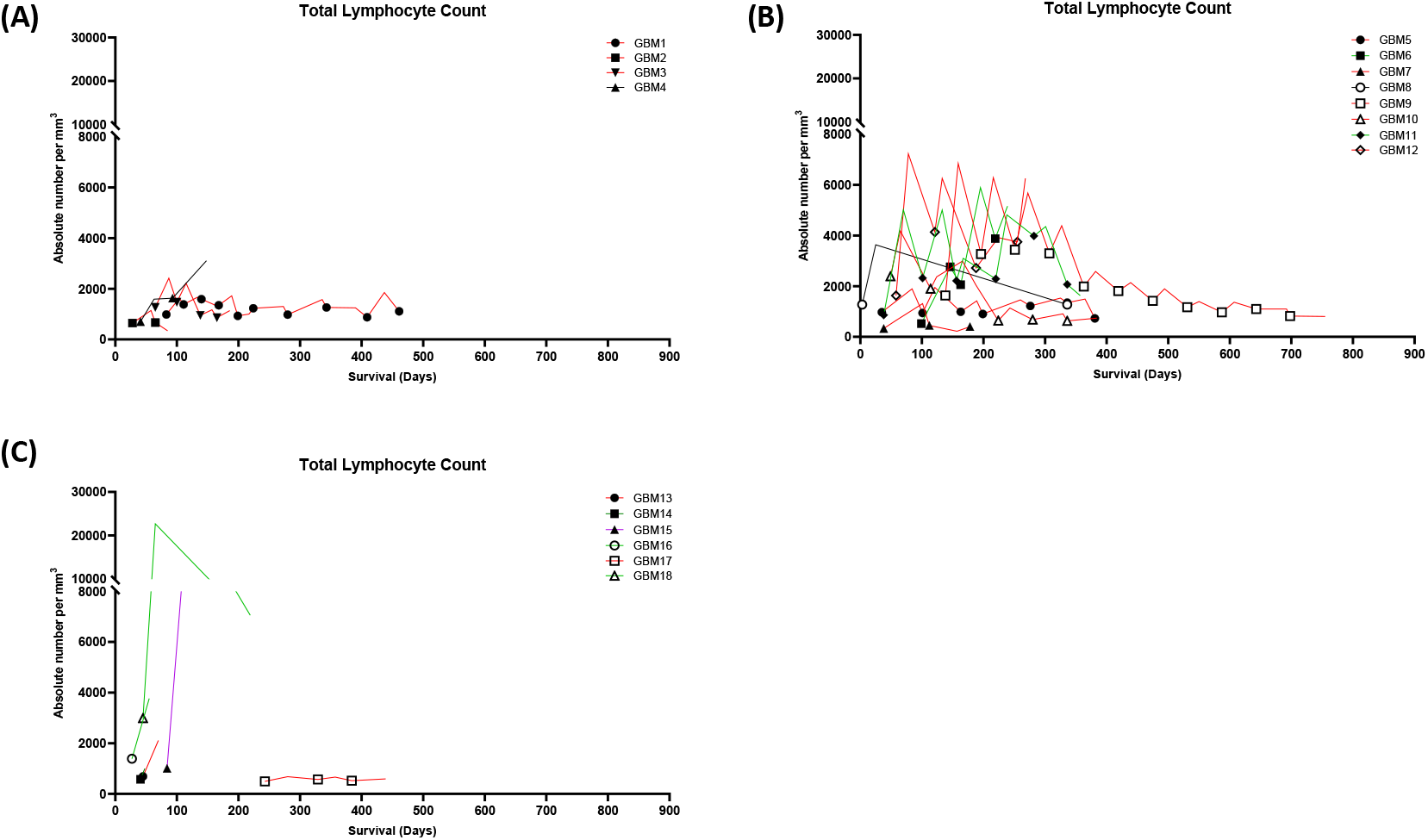
Changes of total lymphocyte counts following treatments of rhIL-7-fyFc. Patients were treated with 120∼240 ug/kg rhIL-7-hyFc (A), 480∼720 ug/kg rhIL-7-hyFc (B) and 960∼1440 ug/kg rhIL-7-hyFc (C). Symbol; rhIL-7-hyFc administration, Red line; TMZ treatment in addition to rhIL-7-hyFc, Green line; Avastin or Avastin+irinotecan treatment in addition to rhIL-7-hyFc, Purple line; PCV treatment in addition to rhIL-7-hyFc.

### Safety profile and survival outcomes

Most common adverse events related to rhIL-7-hyFc were injection site reactions including urticaria, redness, and/or itching sensation at near injection sites (64.7%). Of these patients, only one showed grade 3 urticaria and itching sensation. Other serious adverse events (grade 3-4) observed include edema near injection site (13.2%), fatigue (2.9%), and febrile sensation (2.9%) (Table 3).

**Table 3.**
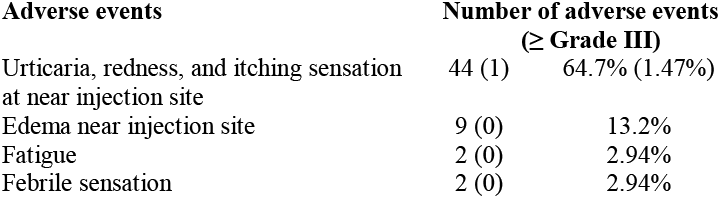
Adverse events to treatment with rhIL-7-hyFc.

Median OS and PFS of enrolled patients after recurrence were 378 days (range 107-864 days) and 231 days (55-726 days), respectively (Figure 3). We also compared the median OS of enrolled patients (treatment group) with that of historical patients with recurrent GBM (historical group) treated in our institution. Median OS of the treatment 18 patients after recurrence was significantly longer than that of 123 patients in the historical cohort (378 days, range 107-864 versus 169 days, range 33-1,067, *p* =0.003) (Figure 3.)

**Figure 3.**
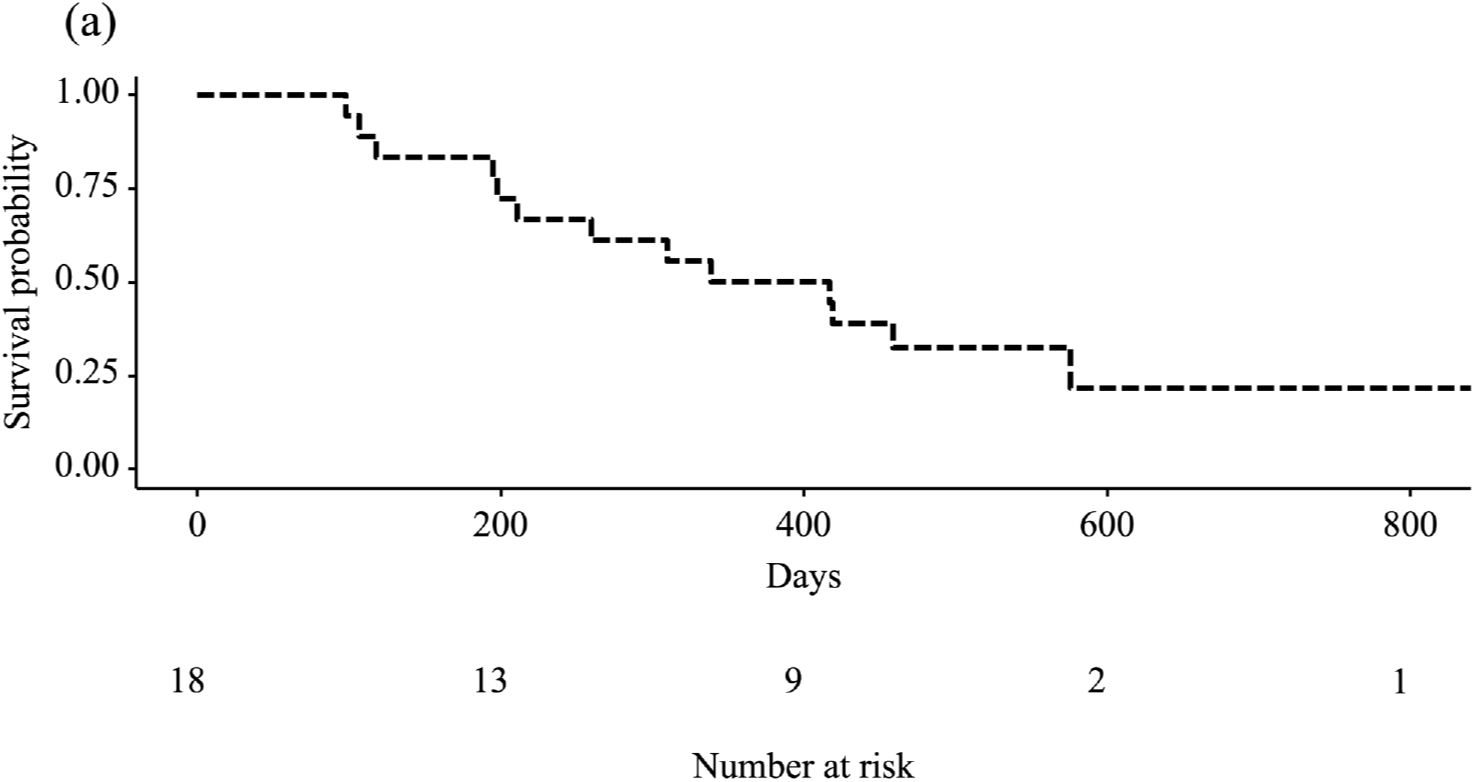
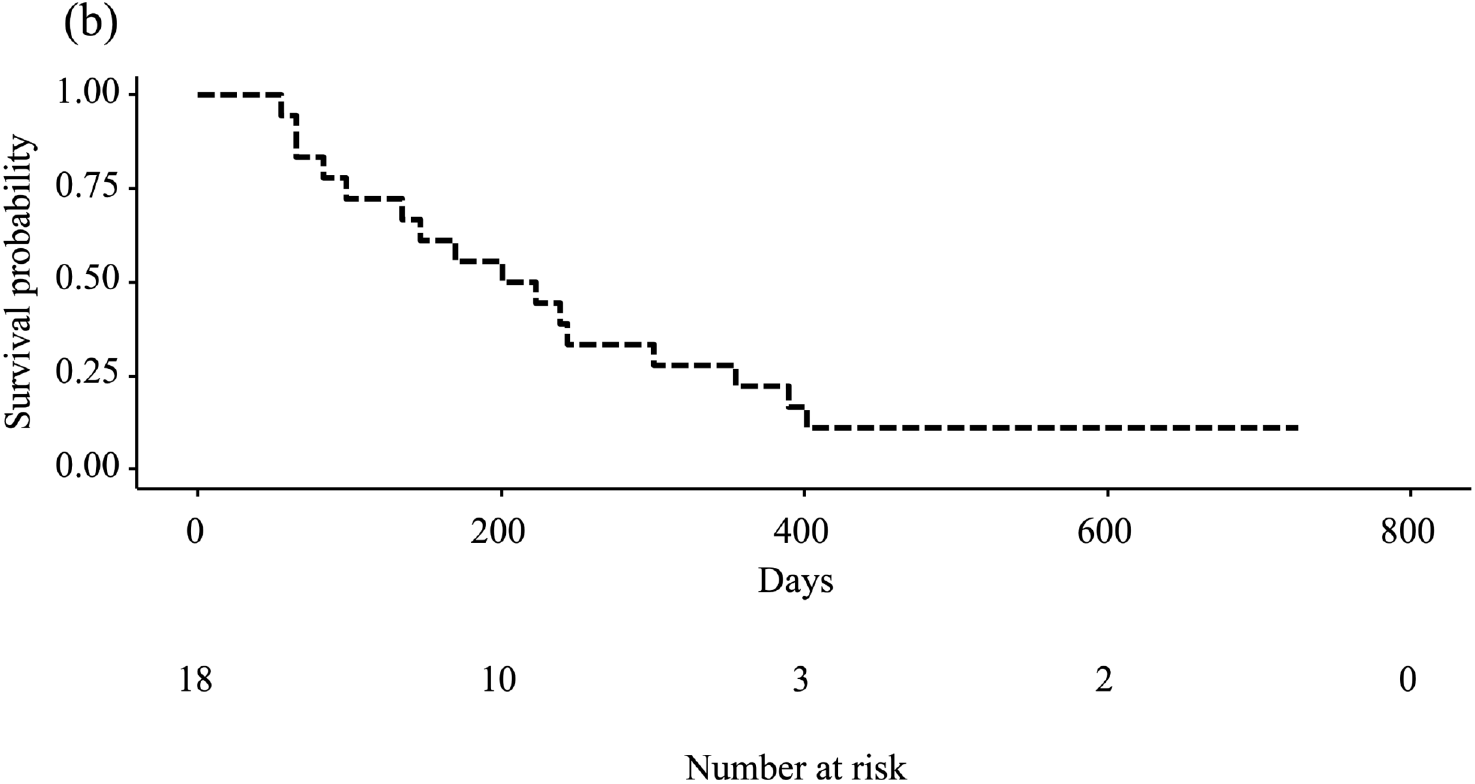
Kaplan-Meier survival curve for overall survival (a) and progression-free survival (b) of enrolled patients.

Of 18 total patients in treatment group, 15 (83.3%) survived more than 6 months and 9 (50.0%) survived more than 1 year after treatment with rhIL-7-hyFc. In addition, 9 patients (50.0%) had stable disease for more than 6 months after co-treatment with rhIL-7-hyFc injection and systemic therapy. Among them, 2 patients (GBM6 & 11) co-treated with bevacizumab had partial response, and one patient (GBM9) co-treated with temozolomide had stable disease for more than 2 years after treatment with rhIL-7-hyFc injection. Representative radiographic findings from these patients are illustrated in Figure 4.

**Figure 4.**
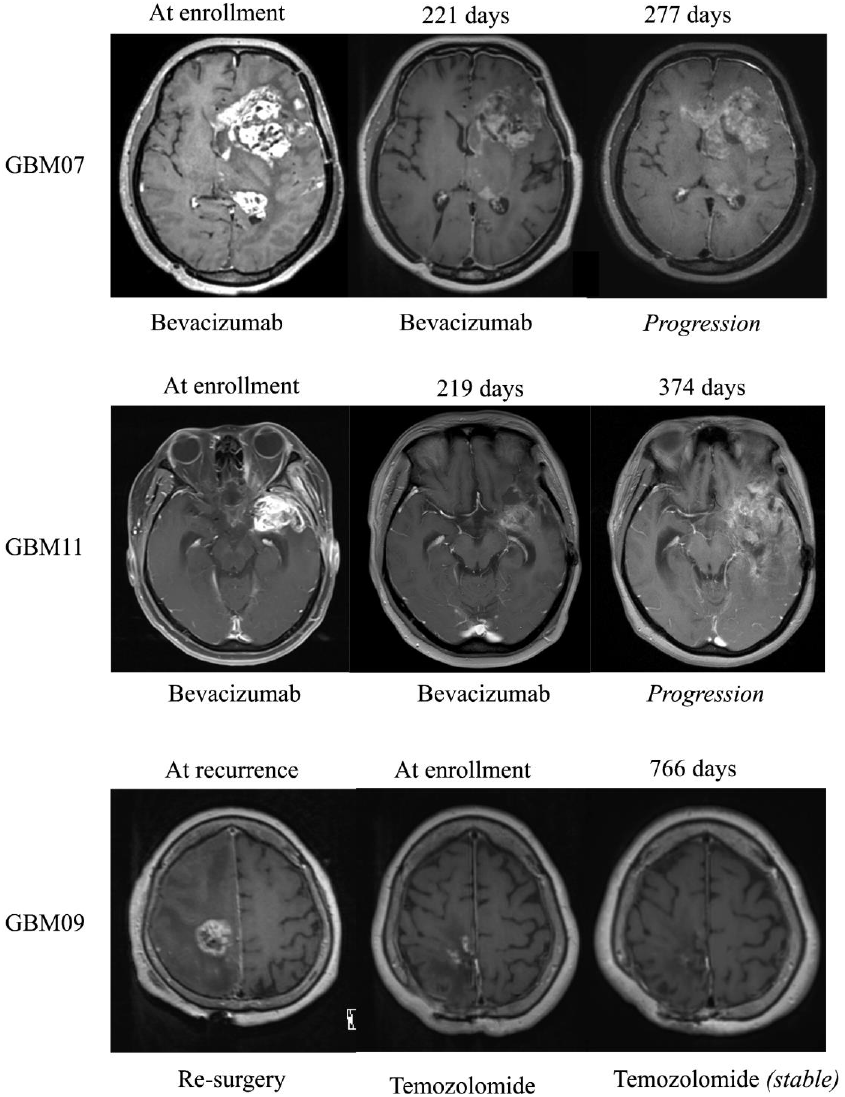
Representative clinical images of patients who showed partial responses or longer stable diseases after injection of rhIL-7-hyFc.

## Discussion

We demonstrated that rhIL-7-hyFc can be safely added to conventional salvageable systemic therapy such as temozolomide, bevacizumab, and PCV chemotherapy. There are concerns whether rhIL-7-hyFc can restore lymphocyte count when used in combination with cytotoxic chemotherapy, but our study showed that TLCs of patients who received co-treatment of rhIL-7-hyFc with chemotherapy were dramatically increased from 1,131 (range 330-2,989) cells/mm^3^ at baseline to 4,356 (range 661-22,661) cells/mm^3^ after the first treatment, a mean 3.40-fold increase (range 1.38-9.13). The increase in TLCs was dose-dependent. In addition, continuing rhIL-7-hyFc therapy facilitated recovery of lymphopenia induced by concurrent chemotherapy and maintained TLCs throughout the treatment higher than the baseline. Mean OS after recurrence of rhIL-7-hyFc treated patients exceeded 1 year (378 days); with 2 patients showing partial response and one patient showing stable disease for more than 2 years. Because of the small number of patients (n=18) and the potential impact of salvageable systemic therapy, it is premature to draw any conclusion, therefore a larger, randomized controlled trial is needed to validate the result. However, our study highlights that rhIL-7-hyFc cytokine therapy effectively and safely restored lymphocyte count and ongoing treatment sustained elevated levels of TLC in spite of concurrent systemic therapy with temozolomide, bevacizumab, or PCV chemotherapy in recurrent GBM patients.

Ample evidence suggests that lymphopenia is one of the poorest prognostic factors in various cancer types including GBM ^5-8,10^. The standard care of GBM includes aggressive concurrent chemoradiation followed by chemotherapy using temozolomide, which causes severe and prolonged lymphopenia in most GBM patients ^11,19^. In this context, preventing or restoring lymphopenia has been suggested as a novel immunotherapeutic strategy ^5,9,10^. IL-7, first discovered in the 1980s, is one of the critical cytokines to elicit T-cell responses to target cancer cells ^13^. It promotes lymphocyte development in the thymus and maintains the homeostasis of naive and memory T cells in the periphery ^20,21^. Furthermore, IL-7 can repair T cell injury in cancer patients and overcome immunosuppressive tumor microenvironment ^11,22^. Several preclinical studies have demonstrated the efficacy of IL-7 in various cancer types with or without combined therapeutic agents ^23-25^.

Rosenberg et al. first reported that IL-7 cytokine therapy administered every 3 days for 8 sessions could increase CD4+ and CD8+ T cells in cancer patients ^15^. In another clinical trial, recombinant human IL-7 injection every other day for 2 weeks for refractory cancer patients increased CD3+, CD4+, and CD8+ lymphocytes in a dose-dependent manner ^16^. In another randomized placebo-controlled phase IIa clinical study, patients with metastatic breast cancer who received 3 injections of recombinant IL-7 for 3 weeks had increased CD4 T cell count compared to patients who received placebo ^14^. Lastly, a recent clinical trial using rh-IL-7 as adjuvant therapy every 2 weeks combined with dendritic cell vaccination or autologous lymphocyte infusion showed significantly better OS in pediatric sarcoma patients compared to that of patients in a historical control cohort ^17^. Compared to previous clinical studies multiple administered short-acting IL-7, our study showed that rhIL-7-hyFc could increase TLC drastically even after a single injection. In addition, we demonstrated that rhIL-7-hyFc therapy could be combined with conventional systemic therapy such as temozolomide, bevacizumab, or PCV chemotherapy. Even for patients receiving lymphodepleting cytotoxic chemotherapy, TLC increased after rhIL-7-hyFc injection. Therefore, combination of rhIL-7-hyFc with conventional systemic therapy such as temozolomide, bevacizumab, or PCV chemotherapy can provide an opportunity for clinical benefit to the patients. To the best of our knowledge, our study is the first to report the clinical experience of rhIL-7-hyFC therapy in GBM patients. Our preliminary clinical findings will facilitate the development as a novel combination immunotherapy strategy for various cancers including GBM. Considering that lymphopenia is usually a result of concurrent chemoradiation, IL-7 administered during or after concurrent chemoradiation might be an alternative to restore or prevent lymphopenia. In addition, increasing lymphocyte counts can augment anti-tumor effect of immune checkpoint inhibitors for the typical immunotherapy non-responsive GBM.^10,26^.

Our study had several limitations. First, the selection bias is imposed on the study as compassionate use is designed without strict conditions of enrollment. Second, the various time interval between the time of recurrence and the first injection of rhIL-7-hyFc could complicate the interpretation of the study. Third, a few patients who recurred from primary high-grade glioma were included, although all patients had pathologically confirmed GBM. Last, our study did not include changes in immune subsets including various T cell populations such as CD3, CD4, CD8 positive T cells, or regulatory T cells. Further prospective studies including immune subsets from peripheral blood and/or tumor tissue are highly desirable.

## Conclusion

Our study first reports that IL-7 immunotherapy could restore lymphocyte count and maintain elevated TLC when administered with systemic therapy in recurrent GBM patients without serious adverse events. Combination of rhIL-7-hyFc with conventional systemic therapy such as temozolomide, bevacizumab, or PCV chemotherapy can provide opportunity of clinical benefit to the patients This outcome warrants further larger and randomized clinical trials to validate the clinical benefits of rIhL-7-hyFc for GBM patients.

## Data Availability

All data produced in the present study are available upon reasonable request to the authors.

---

**Supplementary Table 1.** Baseline characteristics of historical patients with recurrent GBM in our institution.

| n (%) | N =123 |
| --- | --- |
| Sex |  |
| Female | 55 (44.7) |
| Male | 68 (55.3) |
| Mean age (years) | 59.3 ± 13.2 |
| > 65 years | 49 (39.8) |
| Extent of resection |  |
| Gross total resection | 42 (34.1) |
| Subtotal resection | 36 (29.3) |
| Partial resection | 23 (18.7) |
| Biopsy | 22 (17.9) |
| IDH mutation |  |
| Yes | 2 (1.6) |
| No | 64 (52.0) |
| Unknown | 57 (46.3) |
| 1p19q co-deletion |  |
| Yes | 2 (1.6) |
| No | 100 (81.3) |
| Unknown | 21 (17.1) |
| MGMT methylation |  |
| Yes | 47 (38.2) |
| No | 52 (42.3) |
| Unknown | 24 (19.5) |
| Completion of CCRT<br>and adjuvant chemotherapy |  |
| Yes | 60 (48.8) |
| No | 63 (51.2) |

**Supplementary Table 2.** Changes of total lymphocyte counts following treatments.

| Patient Code | Cycle | Dose, $\mu\text{g/kg}$ | WBC (lymphocyte %) at minimum | TLC at minimum | WBC (lymphocyte %) at maximum | TLC at maximum | Fold change |
| --- | --- | --- | --- | --- | --- | --- | --- |
| GBM1 | 1* | 60 | 7260 (13.6%) | 987 | 7680 (21.2) | 1,628 | 1.65 |
|  | 2* | 60 | 6030 (22.9) | 1381 | 5720 (29.0) | 1659 | 1.20 |
|  | 3* | 60 | 5710 (27.8) | 1587 | 4670 (29.3) | 1368 | 0.86 |
|  | 4* | 60 | 5090 (26.5) | 1349 | 5010 (34.3) | 1718 | 1.27 |
|  | 5* | 60 | 4200 (22.1) | 928 | 4150 (24.1) | 1000 | 1.08 |
|  | 6 | 240 | 4840 (25.4) | 1229 | 6800 (19.1) | 1299 | 1.00 |
|  | 7 | 480 | 7770 (12.6) | 979 | 9730 (16.2) | 1576 | 1.61 |
|  | 8 | 240 | 6250 (20.2) | 1263 | 5450 (22.9) | 1248 | 0.99 |
|  | 9 | 600 | 3180 (27.5) | 875 | 18900 (9.8) | 1852 | 2.12 |
| GBM2 | 1* | 120 | 4240 (15.3) | 649 | 7920 (14.4) | 1140 | 1.76 |
|  | 2* | 120 | 4880 (13.7) | 669 | 5030 (6.8) | 342 | 0.51 |
| GBM3 | 1 | 120 | 5290 (24.0) | 1270 | 5520 (43.8) | 2418 | 1.90 |
|  | 2 | 120 | 4410 (33.3) | 1469 | 6250 (35.5) | 2219 | 1.51 |
|  | 3 | 120 | 7030 (13.5) | 949 | 5320 (22.0) | 1170 | 1.23 |
|  | 4 | 240 | 4740 (18.1) | 858 | 4520 (25.2) | 1139 | 1.33 |
| GBM4 | 1 | 120 | 3640 (19.2) | 699 | 4640 (35.7) | 1592 | 2.28 |
|  | 2 | 120 | 4450 (36.6) | 1692 | N/A | N/A | N/A |
|  | 3 | 120 | 8320 (37.3) | 3103 | N/A | N/A | N/A |
| GBM5 | 1 | 240 | 8550 (11.4) | 975 | 9730 (19.5) | 1897 | 1.95 |
|  | 2 | 480 | 4880 (19.6) | 941 | 3510 (55.5) | 1948 | 2.07 |
|  | 3 | 480 | 2610 (37.9) | 989 | 1600 (49.4) | 790 | 0.80 |
|  | 4 | 480 | 2240 (40.2) | 900 | 5410 (27.0) | 1461 | 1.62 |
|  | 5 | 480 | 5410 (22.5) | 1217 | 5750 (26.6) | 1530 | 1.26 |
|  | 6 | 720 | 8000 (16.9) | 1352 | 6080 (24.5) | 1490 | 1.10 |
|  | 7 | 720 | 3560 (20.5) | 730 | N/A | N/A | N/A |
| GBM6 | 1 | 600 | 2590 (20.1) | 521 | 4670 (59.1) | 2760 | 5.30 |
|  | 2 | 600 | 8770 (23.5) | 2061 | 9770 (60.3) | 5891 | 2.86 |
|  | 3 | 600 | 8120 (47.8) | 3881 | 11870 (43.5) | 5163 | 1.33 |
| GBM7 | 1 | 480 | 2000 (16.5) | 330 | 2790 (23.7) | 661 | 2.00 |
|  | 2 | 480 | 4980 (26.3) | 1310 | 1130 (39.8) | 450 | 0.34 |
|  | 3 | 720 | 2470 (8.9) | 220 | 1900 (13.8) | 400 | 1.82 |
| GBM8 | 1 | 720 | 4990 (25.5) | 1272 | 11530 (31.5) | 3632 | 2.85 |
| GBM9 | 1 | 720 | 4690 (34.8) | 1632 | 12310 (55.6) | 6844 | 4.19 |
|  | 2 | 720 | 6900 (47.4) | 3271 | 11860 (53.0) | 6286 | 1.92 |
|  | 3 | 720 | 7810 (44.0) | 3436 | 11140 (51.0) | 5681 | 1.65 |
|  | 4 | 720 | 8120 (40.6) | 3297 | 9730 (45.1) | 4388 | 1.33 |
|  | 5 | 720 | 7180 (27.7) | 1989 | 7900 (32.7) | 2583 | 1.30 |
|  | 6 | 720 | 5420 (33.4) | 1810 | 5650 (37.9) | 2141 | 1.18 |
|  | 7 | 720 | 4640 (30.6) | 1420 | 5800 (32.7) | 1897 | 1.34 |
|  | 8 | 720 | 4860 (24.1) | 1171 | 5200 (26.8) | 1394 | 1.19 |
|  | 9 | 720 | 3140 (30.9) | 970 | 6480 (21.1) | 1367 | 1.41 |
|  | 10 | 720 | 4660 (23.6) | 1100 | 3590 (30.6) | 1099 | 1.00 |
|  | 11 | 720 | 3710 (22.1) | 820 | N/A | N/A | N/A |
|  | 12 | 720 | 4010 (20.0) | 802 | N/A | N/A | N/A |
| GBM10 | 1 | 720 | 8490 (28.2) | 2394 | 9340 (44.8) | 4184 | 1.75 |
|  | 2 | 720 | 9170 (20.7) | 1898 | 4900 (48.4) | 2372 | 1.25 |
|  | 3 | 720 | 7160 (41.8) | 2993 | 5320 (38.3) | 2038 | 0.68 |
|  | 4 | 720 | 6260 (10.2) | 639 | 3390 (33.6) | 1139 | 1.78 |
|  | 5 | 720 | 6650 (10.2) | 678 | 4590 (19.8) | 909 | 1.34 |
|  | 6 | 720 | 5430 (11.6) | 630 | 13660 (5.4) | 738 | 1.17 |
| GBM11 | 1 | 720 | 1850 (46.5) | 860 | 7140 (70.3) | 5019 | 5.83 |
|  | 2 | 720 | 3510 (66.1) | 2320 | 7320 (68.4) | 5007 | 2.16 |
|  | 3 | 720 | 3690 (59.9) | 2210 | 5240 (59.2) | 3102 | 1.40 |
|  | 4 | 720 | 4170 (54.9) | 2289 | 8820 (54.6) | 4816 | 2.10 |
|  | 5 | 720 | 7660 (52.0) | 3983 | 8120 (53.6) | 4352 | 1.09 |
|  | 6 | 720 | 4100 (50.5) | 2071 | 3840 (42.4) | 1628 | 0.79 |
| GBM12 | 1 | 720 | 5920 (27.5) | 1628 | 13260 (54.5) | 7227 | 4.44 |
|  | 2 | 720 | 7850 (52.7) | 4137 | 12740 (49.1) | 6255 | 1.51 |
|  | 3 | 720 | 6920 (39.3) | 2720 | 7000 (56.0) | 3920 | 1.44 |
|  | 4 | 720 | 8410 (44.6) | 3751 | 11790 (53.1) | 6260 | 1.67 |
| GBM13 | 1 | 720 | 4230 (16.3) | 689 | 12550 (16.8) | 2108 | 3.06 |
| GBM14 | 1 | 1,200 | 8130 (7.1) | 577 | 7600 (13.2) | 1003 | 1.74 |
| GBM15 | 1 | 1,200 | 8250 (12.2) | 1007 | 18800 (48.9) | 9193 | 9.13 |
| GBM16 | 1 | 1,440 | 6230 (22.3) | 1389 | 7830 (48.1) | 3766 | 2.71 |
| GBM17 | 1 | 1,200 | 2530 (19.4) | 491 | 2830 (24.0) | 679 | 1.38 |
|  | 2 | 1200 | 3380 (16.9) | 571 | 2880 (22.9) | 660 | 1.15 |
|  | 3 | 1200 | 3120 (16.7) | 521 | 2880 (20.5) | 590 | 1.13 |
| GBM18 | 1 | 1200 | 5860 (51.0) | 2989 | 26850 (84.4) | 22661 | 7.58 |

1 \* *Injected every 4 weeks*

**Figure Suppl.**
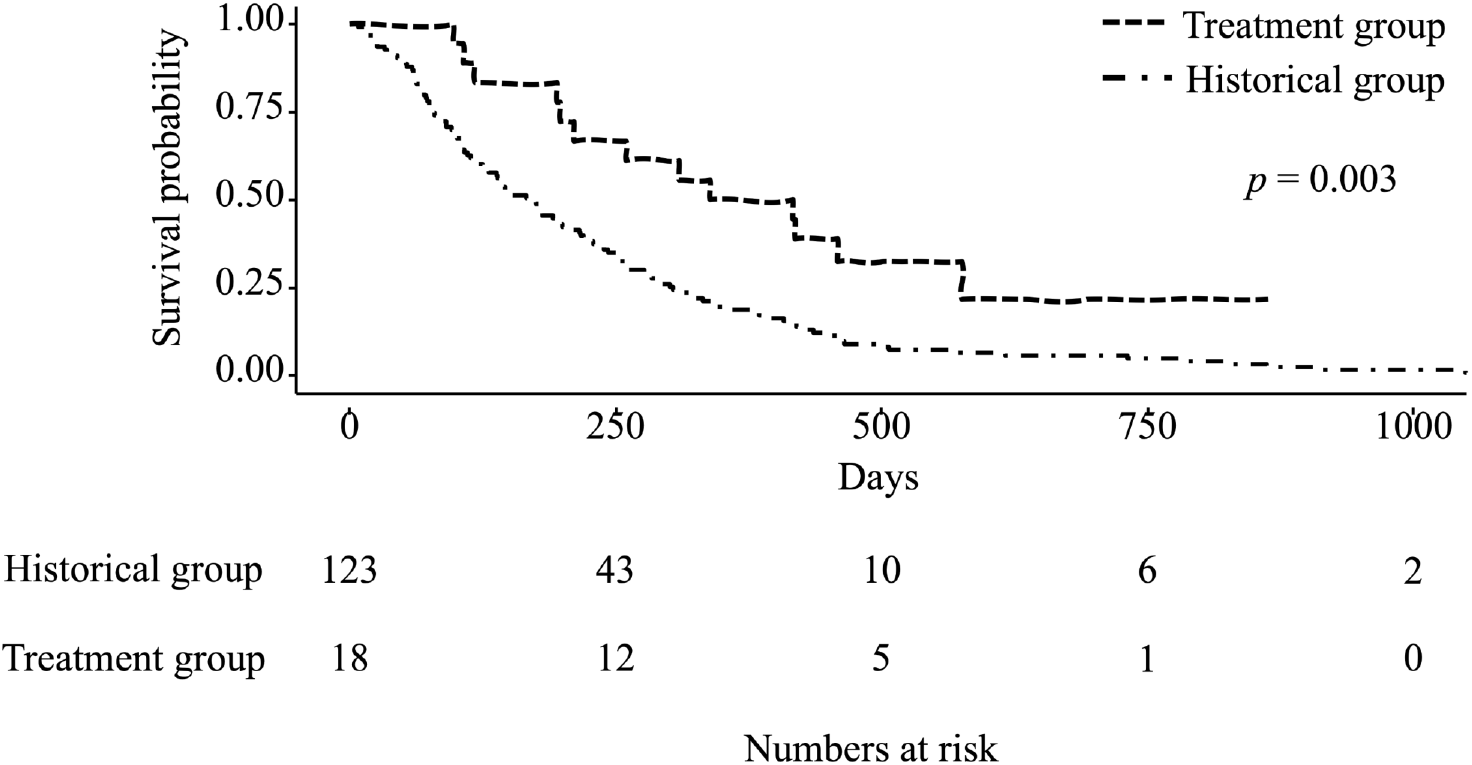
Kaplan-Meier survival curve for overall survival comparing the treatment group with the historical group. The P-value is derived by log-rank test.

